# LGE-MRI for diagnosis of left atrial cardiomyopathy as identified in high-definition endocardial voltage and conduction velocity mapping

**DOI:** 10.1101/2022.02.02.22269817

**Authors:** Deborah Nairn, Martin Eichenlaub, Björn Müller-Edenborn, Heiko Lehrmann, Claudia Nagel, Luca Azzolin, Giorgio Luongo, Rosa M. Figueras Ventura, Barbara Rubio Forcada, Anna Vallès Colomer, Thomas Arentz, Olaf Dössel, Axel Loewe, Amir Jadidi

## Abstract

**Background:** Electro-anatomical voltage, conduction velocity (CV) mapping and late-gadolinium-enhancement-magnetic-resonance-imaging (LGE-MRI) are different diagnostic modalities for atrial cardiomyopathy (ACM). However, discordances remain in the location and extent of detected ACM.

**Objectives:** (1) Comparison of ACM extent and location between current modalities. (2) Development of new estimated optimised image-intensity-thresholds (EOIIT) for LGE-MRI identifying patients with ACM.

**Methods:** Thirty-six ablation-naive persistent AF patients underwent LGE-MRI and high-definition electro-anatomical mapping in sinus rhythm. Significant ACM was defined as low-voltage-substrate (LVS) extent≥5% of the left atrium (LA) surface at <0.5 mV. LGE areas were classified using the Utah, image-intensity-ratio (IIR>1.20) and new EOIIT method for comparison to LVS and slow-conduction-areas <0.2 m/s. ROC-analysis determined the LGE-extent enabling accurate diagnosis of ACM.

**Results:** The degree and distribution of detected pathological substrate varied significantly (p<0.001) across the mapping modalities: 3% (IQR 0-12%) of the LA displayed LVS<0.5 mV vs. 14% (3-25%) slow-conduction-areas<0.2 m/s vs. 16% (6-32%) LGE with Utah method vs. 17%(11-24%) using IIR>1.20, with enhanced discrepancies on posterior LA. A linear correlation was found between the OIIT and each patient’s mean blood pool intensity (R^2^=0.89, p<0.001). LGE-MRI-based ACM-diagnosis improved with the novel EOIIT (83% sensitivity, 88% specificity, AUC:0.94) in comparison to the Utah method (60% sensitivity, 75% specificity, AUC:0.76), and IIR>1.20 (58% sensitivity, 75% specificity, AUC:0.71)

**Conclusion:** Important discordances in distribution of pathological substrate exist between LA-LVS, CV and LGE-MRI, irrespective of the LGE-detection protocol that is used. However, the new EOIIT method improves LGE-MRI-based ACM-diagnosis in ablation-naive AF-patients.

## 1. Introduction

Atrial fibrillation (AF) is the most common cardiac arrhythmia causing an irregular heart rhythm, associated with increased risk for stroke and heart failure [1]. Pulmonary vein isolation (PVI) is a commonly used treatment for AF with a high success rate (75-90%) in paroxysmal AF patients [2]. However, persistent AF patients may present an atrial cardiomyopathy (ACM) with extra-PV pathological substrate, which contributes to the maintenance of AF and reduces the success rate markedly [3, 4].

Electro-anatomical mapping (EAM) during sinus rhythm (SR) identifying low voltage bipolar electrograms (<0.5 mV) is a common technique used to locate pathological substrate [5, 6]. Recently, conduction velocity (CV) mapping has also been investigated due to the structural and functional abnormalities resulting in conduction slowing in the context of AF-associated ACM [7, 8, 9]. Another commonly used method is late gadolinium enhancement magnetic resonance imaging (LGE-MRI) [10, 11, 12]. The advantage of LGE-MRI is that it is a non-invasive diagnostic method. However, the spatial resolution of LGE-MRI is limited. Although all the above-mentioned methods detect pathological substrate, discordances in their location and extent have been reported when mapping during AF [13].

This study assessed the spatial distribution of pathological left atrial substrate as detected in LGE-MRI (using various MRI post-processing methods), voltage and CV mapping during sinus rhythm (SR) in ablation-naive AF patients. The mapping modalities were compared to one another. Additionally, a new estimated optimised image-intensity-thresholding method (EOIIT) for LGE-MRI was developed, enabling identification of patients with atrial cardiomyopathy (ACM) as diagnosed by low-voltage areas at <0.5 mV in SR.

## 2. Methods

### 2.1. Electro-anatomical mapping (EAM)

High-density (>1,200 mapped sites per left atrium (LA)) activation and voltage mapping was performed using a 20-polar Lasso-Nav mapping catheter or a PentaRay catheter (electrode size: 1 mm, spacing: 2–6–2 mm). Mapping was conducted while the patient was in SR prior to PVI using the CARTO-3 mapping system (Biosense Webster, Diamond Bar, CA, USA). Further details about the signal processing and calculation of the LAT, voltage and CV can be found in the supplementary material section 1.1.

In two series of analyses, cut-off values of <0.5 and <1.0 mV were applied to the bipolar voltage maps to define the low voltage substrate (LVS) [14, 15]. Additionally, a CV cut-off value was set at 0.2 m/s to indicate pathological slow conducting substrate [16].

### 2.2. Late gadolinium enhancement magnetic resonance imaging (LGE-MRI)

LGE-MRI was performed on a 3 T scanner (Somatom Skyra, Siemens Healthcare, Erlangen, Germany) as described previously [10, 11]. In brief, LGE-MRI was acquired 15 minutes after contrast injection with a dose of 0.1 mmol Gadoteridol per kg body weight (ProHance®, Bracco, Milan, Italy). Voxel size was 1.25×1.25×2.5 mm (reconstructed to 0.625×0.625×1.25 mm), repetition time was 3.1 ms, echo time was 1.4 ms and flip angle was 14^°^.

Two independent expert core laboratories performed the LA segmentation and detection of LGE-areas: Merisight (Marrek Inc., Sandy, Utah, USA) for the Utah method and the Adas group (Adas3D Medical SL, Barcelona, Spain) for the “image-intensity-ratio (IIR)” method by Benito et al. [12]. All image analysts were blinded to any clinical data. Further details about the two methods can be found in the supplementary material section 1.2.

### 2.3. Analysis

#### 2.3.1. LA mean geometry and statistical analysis of pathological substrate

For this study, geometries obtained from each modality for all patients were aligned to a mean LA shape. Thus, a direct comparison between the EAM and LGE-MRI and between patients could be performed without variations caused by spatial displacement. The steps behind this approach are introduced in the supplementary material, section 1.2 and figure S1. Information on the pulmonary veins (PVs) and mitral valve (MV) were excluded from all substrate analyses.

The median value for each point on the geometry across all patients was calculated for the voltage, CV and MRI maps (image-intensity values), providing a visual comparison between mapping modalities without the influence of outliers due to patient-specific differences.

A spatial histogram for each mapping modality was created to assess which areas most commonly exhibit pathological substrate. Providing for the first time a spatial histogram of LVS and slow conduction areas and a comparison of LGE-MRI spatial histograms with the one previously reported by Higuchi et al. [17].

Additionally, a correlation analysis was performed between each pair of mapping modalities identifying the difference in pathological substrate extent for each patient.

#### 2.3.2. Detection of LA LGE-areas based on estimated optimised image intensity thresholding method “EOIIT”

For this analysis, the individual patient’s “optimised image-intensity-threshold” was considered as the threshold with best quantitative match between LGE-extent and LVS-extent (<0.5 mV) and plotted against the mean bloodpool value of the LA-LGE-MRI for each patient. A linear correlation was calculated using a leave-one-out cross-validation method to determine the EOIIT method for the entire cohort. This analysis was performed for the (1) entire LA, (2) anterior LA and (3) posterior LA. Details on this semi-automatic segmentation approach are given in the supplementary material section 1.3.

The performance of the new thresholds were then evaluated by examining the relationship between the LVS-extent (<0.5 mV) and LGE-MRI substrate for each MRI post-processing method (Utah, IIR>1.20 and EOIIT). Patients with a LVS-extent ≥5 cm2 of LA surface corresponding to a LVS-extent of ≥5% at <0.5 mV were classified as having substantial extent of LVS [18].

Reciever operating curves were created for the IIR>1.2 method and the EOIIT method for the entire LA, anterior and posterior. In this way, the most sensitive and specific LGE-extent was determined to detect significant LA-LVS. A fibrotic tissue extent ≥20% of the total LA-wall (Utah-stages III and IV) was classified as relevant LGE for the Utah method [19].

## 3. Results

### 3.1. Patient characteristics

Thirty-six patients with persistent AF (66±9 years old, 84% male) underwent EAM and LGE-MRI prior to PVI. All patients were electrically cardioverted 4-6 weeks prior to PVI. Fourteen (39%) patients with AF recurrence were cardioverted again on admission. The institutional review board approved the study and all patients provided written informed consent prior to enrollment. The supplementary material provides details of the patients’ characteristics (Table S1) and EAM and LGE-MRI data (Table S2).

### 3.2. Spatial distribution of left atrial pathological substrate in electro-anatomical voltage-activation mapping versus LGE-MRI

Figure 1 reveals discrepancies in the distribution and extent of pathological substrate between EAM modalities and LGE-MRI. Both LGE-MRI methods identified extensive pathological substrate on the inferior wall, whereas the electrograms in this area (marked with a square) are non-fractionated signals with high amplitudes. In contrast, low amplitude signals are seen (marked with a circle) where LVS and slow CV were identified. All patient-specific substrate maps are shown in the supplementary material figure S2.

**Figure 1:**
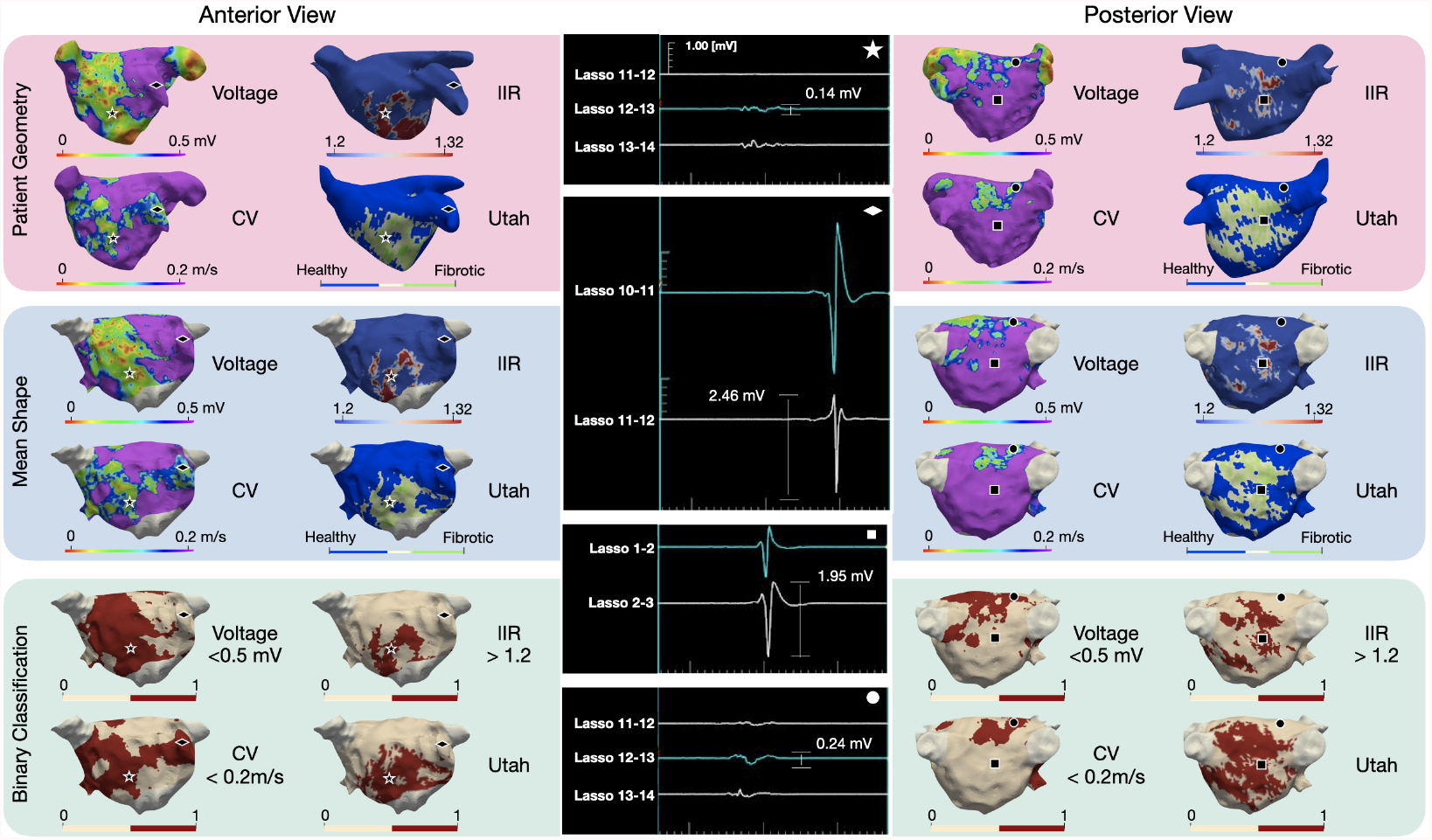
Three-dimensional distribution of voltage <0.5 mV, CV <0.2 m/s and LGE-areas detected with IIR 1.2-1.32 and the Utah method in a representative patient. The top and middle row show the EAM and LGE-MRI information on the patient’s geometry and the mean shape, respectively. The bottom row shows a binary classification, where pathological substrate is shown in red and healthy tissue in cream. Each geometric shape represents a point on the map where the corresponding electrogram is shown in the middle column.

### 3.3. Spatial distribution of left atrial pathological substrate as identified by median voltage values, conduction velocity and LGE-MRI intensity

Figure 2 reveals notable differences in the median voltages between the anterior (median 1.22 mV, IQR 1.05-1.46 mV) and posterior wall (median 1.58 mV, IQR 1.34-1.90 mV). The CV shows similar differences although less pronounced (anterior: median 0.27 m/s, IQR 0.25-0.29 m/s and posterior: median 0.33 m/s, IQR 0.31-0.35 m/s). On the other hand, the LGE-MRI IIR method reveal relatively little difference between the anterior and posterior wall, with median IIR values 0.98 (IQ3 0.96-1.11) and 0.99 (IQ3 0.90-1.12), respectively, signifying more pathologically enhanced tissue at posterior LA.

**Figure 2:**
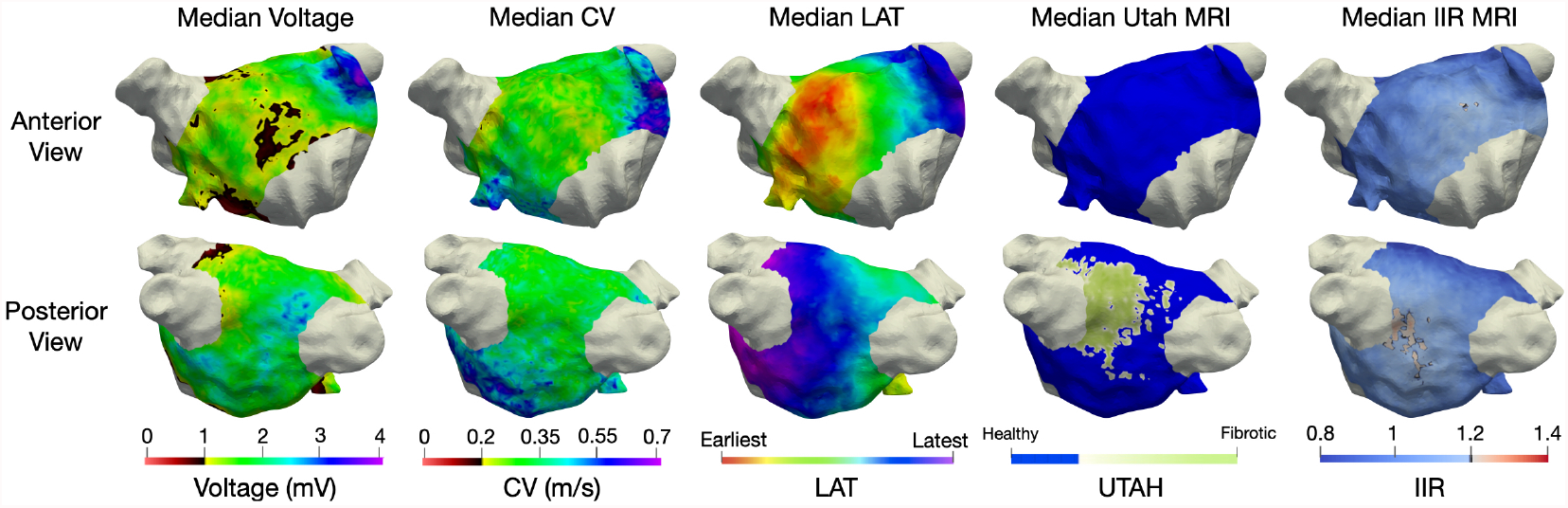
Spatial distribution of the pathological substrate as detected by the different mapping modalities. Each column shows the median values of total study cohort on a representative LA geometry of the mapping modality mentioned at the head of the column.

### 3.4. Spatial localisation frequency of atrial sites displaying pathological substrate

Figure 3 shows that the highest probability for LGE-MRI areas was found around the left inferior PV (with the UTAH method median 56% and IIR>1.20 44% of the patients). In contrast, the LVS <0.5 mV spatial histogram rarely (8% of patients) indicated pathological substrate on posterior wall. Evaluating LVS <1 mV and CV <0.2 mV, the percentage of patients with pathological substrate for both methods was 28% adjacent to the left inferior PV. All methods identified a similar percentage of patients with pathological substrate on the anterior wall (22% LVS <0.5 mV, 22% IIR>1.2, 25% UTAH, 31% CV< 0.2 m/s, 42% LVS < 1 mV).

**Figure 3:**
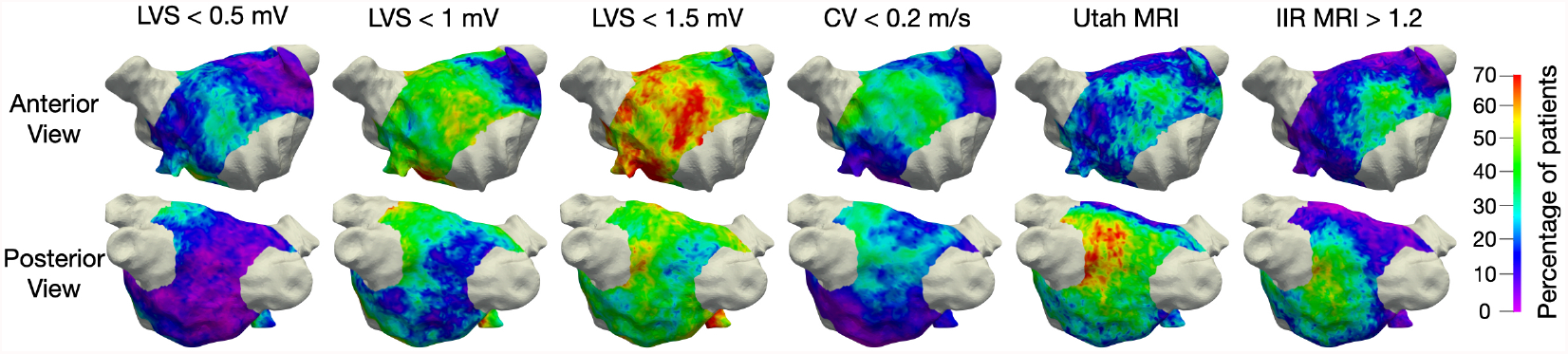
Spatial histogram of left atrial pathological substrate. The colour indicates the percentage of patients for which the respective method identified pathological substrate at the specific location. Each column shows the spatial histogram of the mapping modality mentioned at the top.

### 3.5. Quantification of left atrial pathological substrate

Figure 4 reveals that the lowest difference in terms of global pathological LA substrate extent was found for LVS <0.5 mV and CV <0.2 m/s (median difference 6%) and LGE-extent assessed with IIR >1.2 and UTAH (11%). When comparing LVS <0.5 mV and the LGE-extent with UTAH method, the difference of globally detected substrate was relatively low (median difference 10% of the LA surface; range from 0 to 25%). While both methods detected a similar total extent of pathological LA substrate, the location of detected substrate differed importantly between the methods (see supplementary material figure S2).

**Figure 4:**
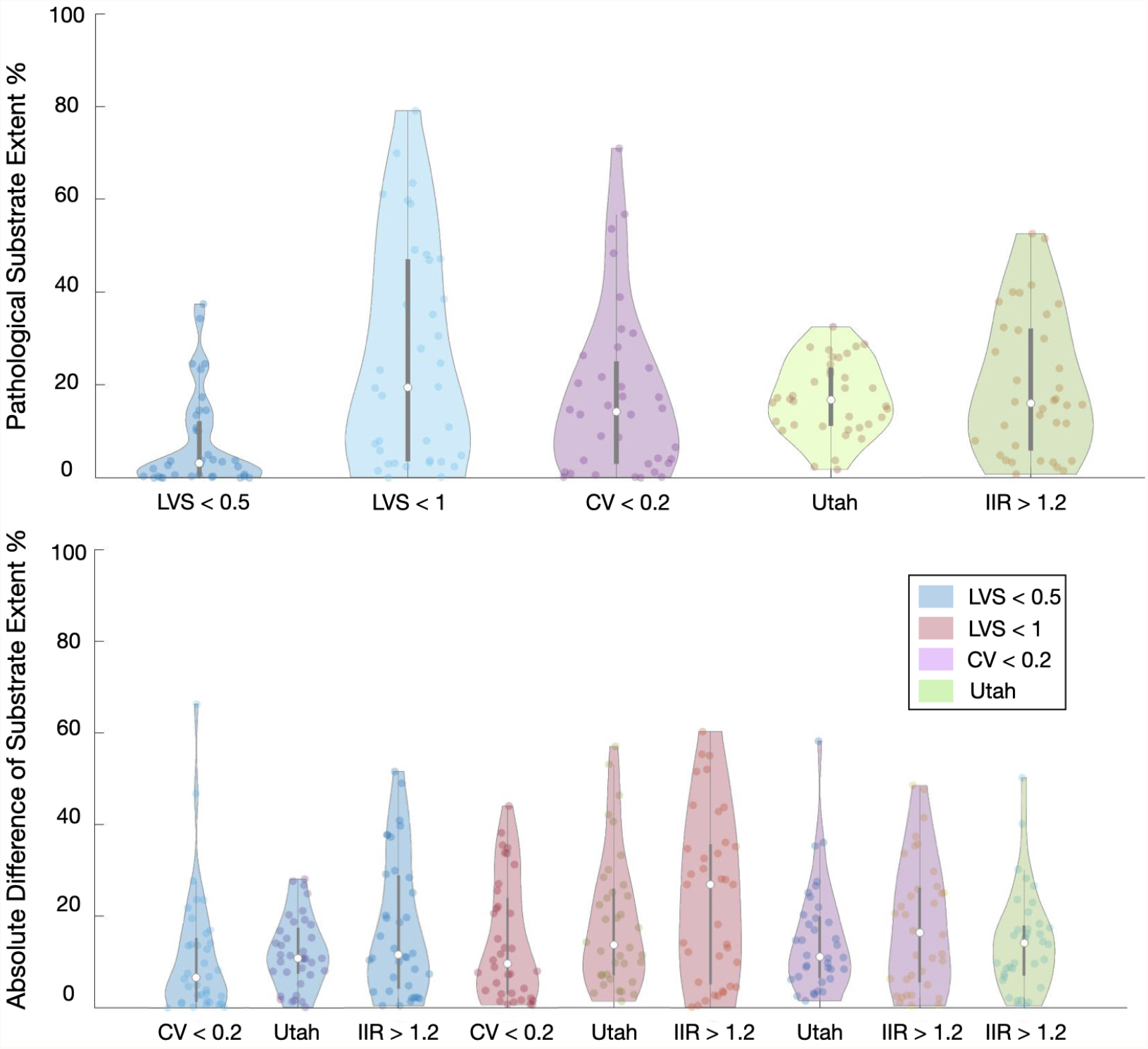
Violin plot demonstrating the extent of pathological substrate (as percentage of LA surface area) for different mapping modalities and the difference of detected substrate between the methods. Top row shows the extent for each mapping modality defined on the x-axis. Bottom row shows the difference in extent of detected substrate for the method on the x-axis with the one identified by the colour of the violin plot as reference. Each dot inside a violin plot represents the value for one patient, the white dot represents the median and the bar shows the interquartile range.

### 3.6. Optimising the LGE-MRI threshold to identify low-voltage substrate

The “optimised-image-intensity-threshold” (with best match in LGE-extent and LVS-extent in an individual patient) was reported to each patient’s left atrial mean bloodpool intensity value (figure 5). Separate analyses were performed on the anterior and posterior LA regions to counter the clear distinction between the two mapping modalities on the posterior wall. For the entire LA, anterior and posterior wall, the linear relationship can be described as *y* = 0.97*x* + 86 (R^2^ = 0.88), *y* = 0.96*x* + 72 (R^2^ = 0.89) and *y* = 0.93*x* + 85 (R^2^ = 0.86), respectively, corresponding to the estimated optimised-image-intensity-threshold (EOIIT). Depending on the individual patient’s mean bloodpool intensity value, the EOIITvalue can be determined using the linear correlation, improving the concordance between LGE-extent and LVS-extent (figure 6CD).

**Figure 5:**
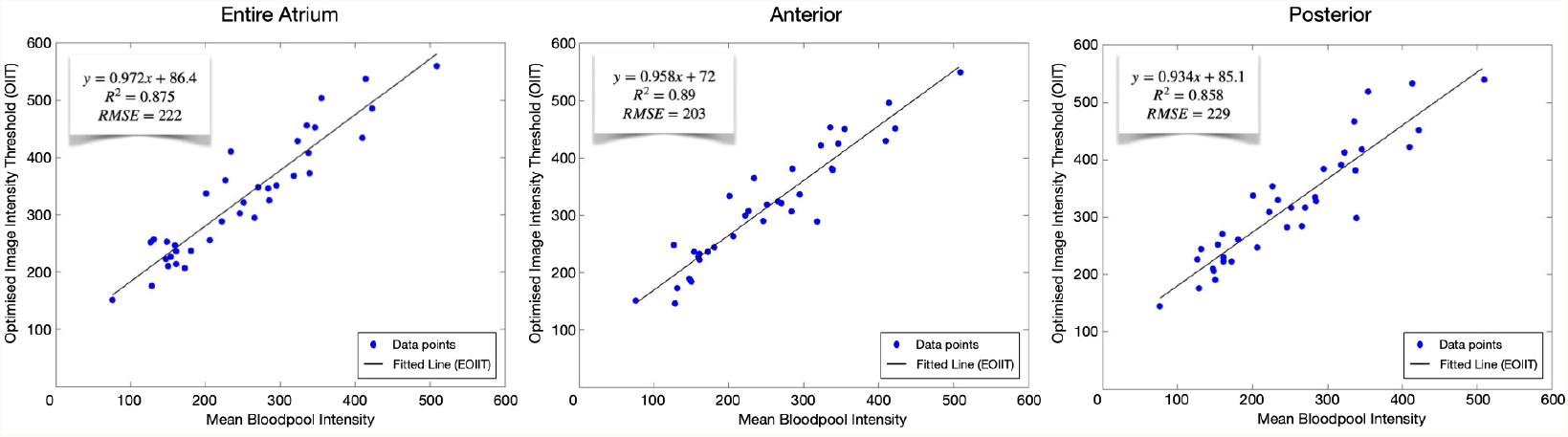
Identification of the EOIIT using linear correlation between the individual patient’s mean bloodpool intensity (x-axis) and optimal image-intensity-threshold (y-axis). From left to right, the results for the entire left atria, anterior wall and posterior wall are shown. Each blue dot represents one patient and the black line is the best linear fit.

**Figure 6:**
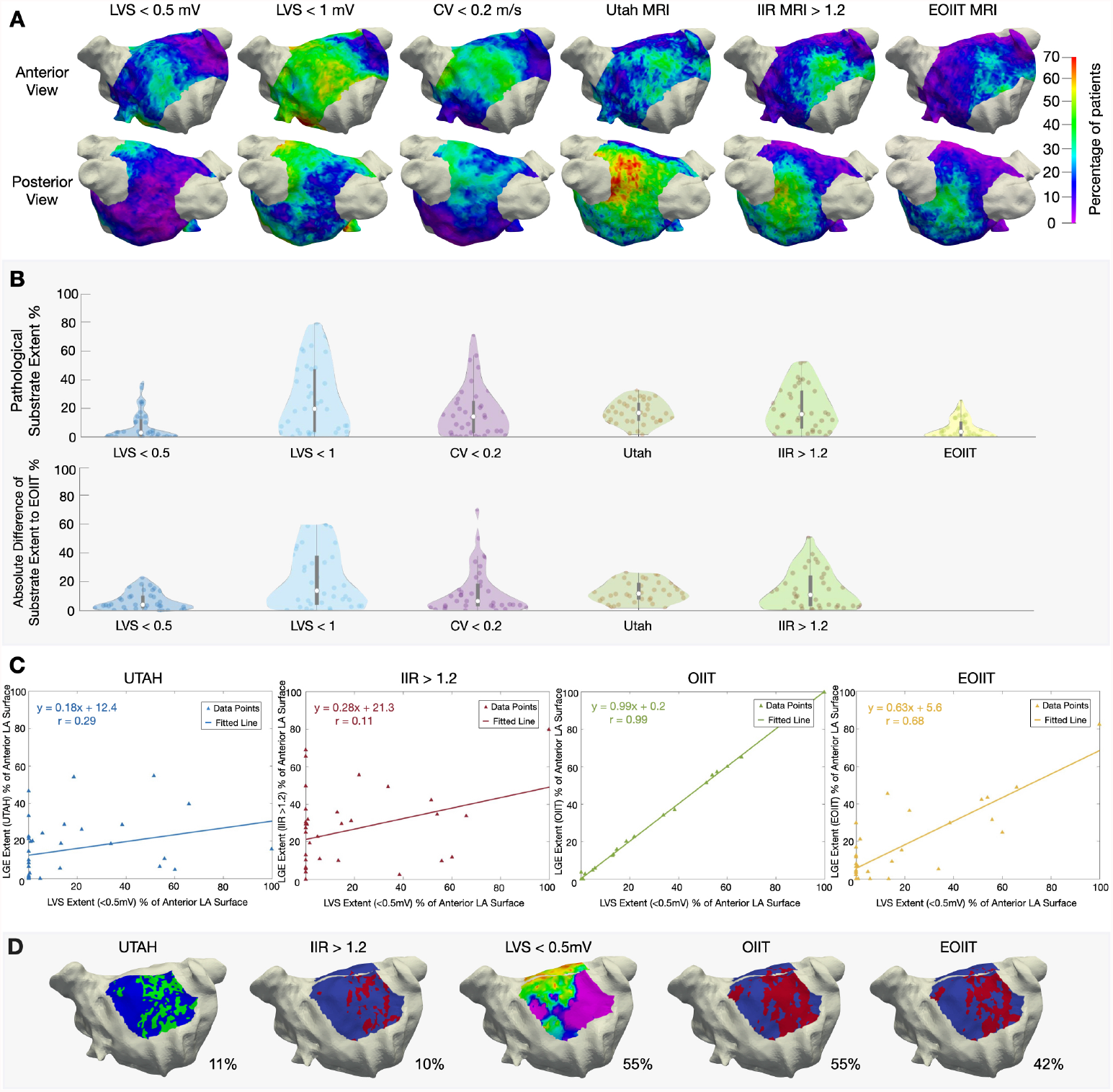
(A) Spatial histogram of each mapping modality including the new EOIIT method. (B) violin plot showing the pathological substrate extent for different mapping modalities and the difference between them and the new EOIIT method. (C) Percentage of substrate extent of each LGE-MRI method versus LVS-extent for the anterior LA-wall. (D) Distribution extent of LVS and LGE-areas for the anterior LA in patient 35.

Supplementary material figure S3 demonstrates the distribution extent of LVS and LGE-areas (for the entire LA, anterior and posterior LA) for both the IIR>1.2 and the new EOIIT method for all patients. Analysis of EOIIT-based LGE-extent revealed similarities with LVS-extent (figure 6A and B) with good correlation on the anterior wall (r=0.68, figure 6C). Supplementary material section 2.4 provides more information on the performance of the new EOIIT compared to voltage on the posterior wall, the entire atrium and CV mapping. Detection of LA-LGE-areas using the new EOIIT-method still did not provide an exact match to LVS-extent in every patient. This is due to the OIIT-value of each patient not lying precisely on the EOIIT-line (figure 5). As seen in figure 6, EOIIT-based LGE-extent (42%) is similar to LVS-extent<0.5 mV (55%) in patient 35. However, the locations of detected substrate remain different between voltage mapping and LGE-MRI.

### 3.7. Estimated optimised image intensity threshold (EOIIT) enables LGE-MRI-based diagnosis of patients with atrial cardiomyopathy presenting low-voltage substrate

Despite difficulties in exact quantitative match between EOIIT-based LGE-extent and LVS-extent, we hypothesized that the new EOIIT-method could be useful to diagnose patients with significant ACM (defined as LVS <0.5 mV at >5% of LA surface area). ROC-analysis determined the best quantitative LGE-extent for ACM-diagnosis (figure 7). ACM-diagnosis was most accurate when using the EOIIT-based LGE-extent>13% at the anterior LA (sensitivity: 83%, specificity: 88%, AUC: 0.94), compared to IIR>1.20-based LGE-extent≥31% (sensitivity: 58%, specificity: 75%, AUC: 0.71). Figure 8 additionally illustrates these results, where the mid panel dashed line corresponds to >13% EOIIT-based LGE-extent and >31% IIR>1.20-based LGE-extent at the anterior LA. For the entire LA, the EOIIT-based (>4%) and Utah-based (≥20%) LGE-extent had a sensitivity and specificity of 75% and 67% (AUC: 0.65) and 60% and 75% (AUC: 0.76), respectively.

**Figure 7:**
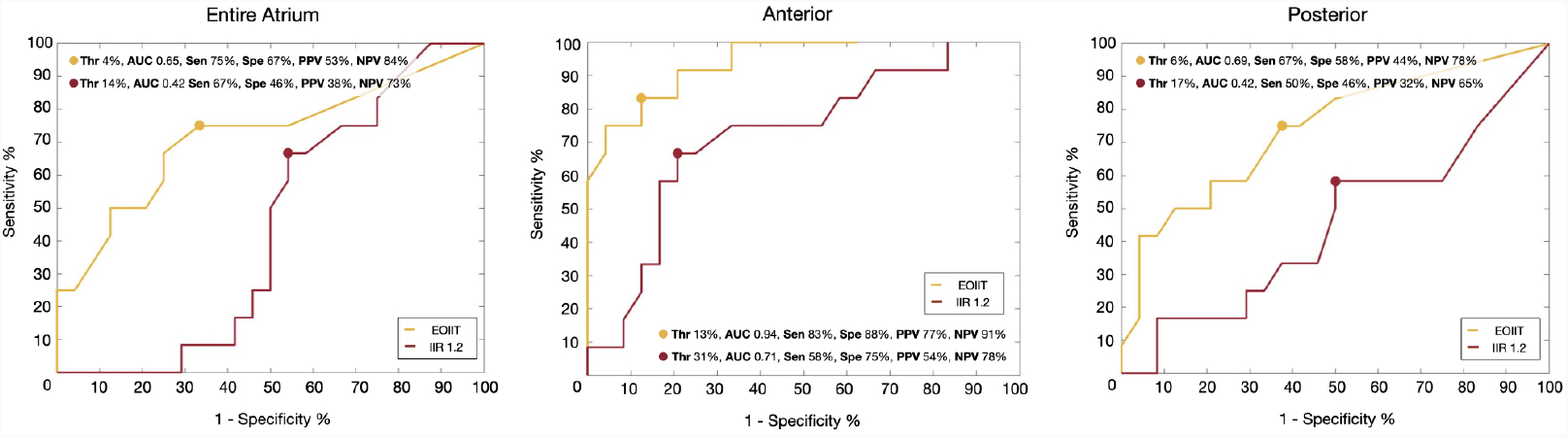
ROC analysis determines the diagnostic LGE-extent that identifies patients with ACM (defined as LVS <0.5 mV at ≥5% of total LA surface. From left to right: results for identification of ≥5% LVS-extent on the entire atria when using LGE-extent from the entire LA, anterior and posterior wall. Yellow and red lines show the ROC of the EOIIT-based and IIR >1.2-based methods for LGE-detection, respectively.

**Figure 8:**
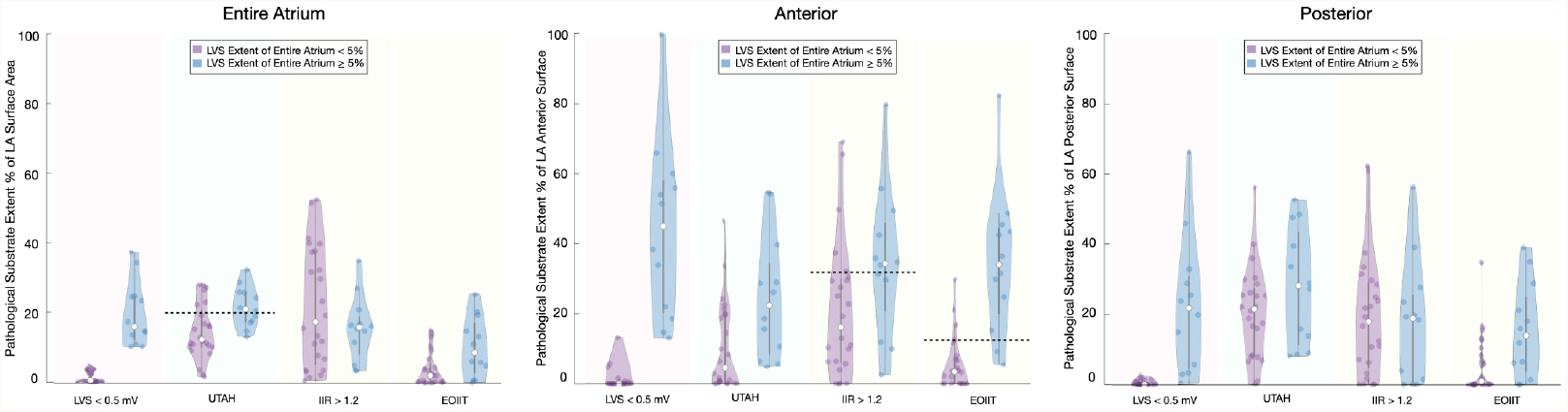
Violin plots illustrate the extent of detected LA substrate compared to pathological low-voltage substrate<0.5 mV at ≥5% of total LA surface. The results for the entire LA, anterior wall and posterior wall are shown from the left to right. The blue violins illustrate the extent of pathological substrate in patients who had significant low-voltage extent ≥5%. The purple violins illustrate the extent of pathological substrate in patients who had no significant low-voltage substrate (low-voltage extent <5%).

## 4. Discussion

### 4.1. Main findings

This study investigated the similarity of different mapping modalities in detecting left atrial pathological substrate. Three key findings can be reported:

1. Important discordances exist in the extent and spatial localisation of identified pathological left atrial substrate when comparing electro-anatomical voltage and conduction velocity mapping to LGE-MRI. These discordances exist with all LGE-detection protocols and are more pronounced on the posterior LA wall.

2. The extent of left atrial slow-conduction-areas (CV<0.2 m/s) correlated moderately (r = 0.57) well to LVS (LVS<0.5 mV).

3. Applying the new EOIIT-method on the anterior LA-wall enables LGE-MRI-based diagnosis of AF patients with ACM (presenting significant low voltage substrate<0.5 mV at ≥5% LA surface) with a sensitivity of 83% and a specificity of 88%. However, significant differences in localisation of LGE-areas to LVS still remain.

### 4.2. Clinical significance of left atrial cardiomyopathy in patients with atrial fibrillation

Several studies have demonstrated the relevance of LA-LVS<0.5mV regarding increased arrhythmia recurrences within 12 months following pulmonary vein isolation (PVI) for AF [14, 20]. Recently, the predictive value of LVS<0.5mV at >5% of LA surface was demonstrated with regard to arrhythmia recurrence rate following PVI. [18]. Therefore, this LVS-extent was used in the current study. In addition, the presence of ACM, as assessed by LA-LVS is associated with an increased risk for ischemic stroke [21]. Therefore, the most recent European guidelines on the management of patients with AF recommend diagnosis of ACM in order to initiate therapeutic efforts in limiting progression and related complications of ACM [1].

### 4.3. Regional differences in detected left atrial substrate - comparing EAM to LGE-MRI

Pathological substrate was most consistently (20-40% of patients) found with all modalities (low-voltage, slow conduction and LGE-MRI) at the anterior LA-wall, confirming preferential profibrotic remodeling in this area. Across the whole LA, the CV was 0.48 ± 0.39 m/s, which is comparable to previous findings in AF patients [8, 9]. Our study confirms that low-voltage and slow-conduction areas most frequently develop on the anteroseptal LA-wall and roof [22, 23].

In contrast, both the Utah- and IIR>1.20 methods most frequently detect LGE at the left posterior LA adjacent to the descending aorta, which is consistent with previous reports [17, 24, 13]. Caixal et al. demonstrated that the spatial proximity of posterior LA-wall to the descending aorta determines the degree of LGE in this region [24]. In a subset of 16 patients, these authors reported reduction of bipolar voltages and CV values in LGE-areas at left posterior LA. In the current study, 56% (Utah-method) and 44% (IIR>1.20-method) of patients presented LGE at posterior-LA adjacent to the left PV, whereas CV<0.2 m/s was found in 28% and LVS<0.5mV in 8% of patients. Although, voltage reduction can be observed in some patients within this LA-area, the electrograms are non-fractionated, and display voltages>1.0 mV in >70% of patients. Thus discrepancies in substrate qualification remain, even when defining LVS as <1.0 mV or <1.5 mV (figure 3). The current study demonstrates that extent and location of LGE-areas differ importantly from LA-LVS<0.5 mV.

### 4.4. Complementarity of mapping systems

Pathological substrate is frequently in opposite locations when comparing electrophysiological substrate to LGE-areas. When considering all patients (supplementary material figures S2-S3), the distribution and location of the pathological substrate is well correlated in some, while they differ in others.

One could hypothesise that the mapping modalities provide complementary information that could be leveraged when determining ablation targets. It has been reported that different fibrotic patterns of various types and densities exist, which could affect the mapping modalities differently [25]. Small patchy fibrotic strands with small contributions to the signal may be compensated by surrounding myocytes and atrial far-fields, causing “undersensing” of fibrotic areas. Additionally, endocardial mapping measurements are dominated by the first 1-2 mm endocardial layer substrate. Fibrosis solely expressed on the epicardial side may be hidden to endocardial catheters. Examining all three methods (CV, LVS and LGE-MRI) may help to comprehensively assess the pathological LA substrate with a more localised analysis than possible with the ECG [26, 27]. However, the fact that ablation of LGE-areas in the DECAAF II study did not improve sinus rhythm maintenance rates, indicates that the detected LGE-areas (using the Utah-method) may not have a major arrhythmogenic role in human AF [19].

### 4.5. Optimising LGE-MRI thresholds

Identifying the same regions of pathological substrate across all established mapping modalities was not possible with the current techniques. Therefore, we sought to improve LGE-based detection of total LA LVS-extent, thus allowing a more accurate ACM-diagnosis. In this analysis, new LGE-MRI thresholds were identified, which, similarly to the IIR method, can be determined using each patient’s mean bloodpool intensity.

Benito et al. studied 30 patients with various stages of AF. They set the thresh-old to determine pathological substrate at >1.20 [12]. As discussed, determining the exact extent of pathological substrate on the thin posterior wall remains challenging, which may be related to MRI resolution [28]. Since the high-intensity areas were mostly located on the posterior wall across the patient cohort, taking the mean tissue IIR + 2 SD as the threshold in the study by Benito et al. was potentially influenced by these high-intensity areas on the posterior wall. Therefore, their suggested threshold might be too high to identify pathological substrate on the anterior wall.

The new EOIIT-based thresholds presented in this work, however, can provide a better estimate of the LVS-extent located on the anterior LA-wall, thus enabling the diagnosis of ACM in individuals. Previous studies revealed that LVS first develops on the anterior LA, and in later stages involves the posterior LA-wall. [22, 23] Therefore, diagnosis of ACM that is based on analysis of LGE-presence on the anterior LA-wall is both sensitive and specific for ACM-diagnosis, as shown in our study. Accurate diagnosis of ACM provides a deeper insight into whether PVI-only would be sufficient or if additional LA mapping is needed.

## 5. Limitations

The CV calculation method in the current study did not exclude uncertain LAT values coming from fractionated multicomponent electrograms, potentially limiting the accuracy of CV-maps. LGE-MRI is limited in its spatial resolution (1-2mm) and cardiac and respiratory motions and adjacent enhanced vascular structures (eg aorta) may produce partial volume effect, resulting in “false” LA-LGE-sites and a lack of spatial correlation between LGE-areas and LVS. Additionally, the patient cohort size for this study was relatively small. Thus, overfitting may play a role when computing the linear fit to determine the new LGE-MRI thresholds. To compensate for this effect, leave-one-out-cross validation was used to obtain the best fit. The new EOIIT-LGE-MRI-based ACM-diagnosis needs further validation in larger external cohorts, assessing its predictive value for arrhythmia recurrence following PVI for persistent AF. Furthermore, adaptations for determination of lab-specific EOIIT-curves may be necessary due to the fact that LGE-detection depends on multiple factors (e.g. type, dosage and timing of contrast injection, magnetic field strength, acquisition protocol).

## 6. Conclusion

Important discordances exist in localisation of detected pathological LA-LVS vs. slow conduction vs. LGE-areas in MRI. Despite a good correlation between the extent of LA-LVS and EOIIT-LGE-areas, the exact locations of identified pathological substrate differ. The new EOIIT-method enables more accurate LGE-MRI-based ACM diagnosis in ablation-naive AF-patients.

## Data Availability

To protect the safety of the patients, the data used for this study can not be provided. However, the figures within the article and supplementary material show detailed analyses for all patients used. Requests to access these datasets should be directed to: Deborah Nairn.

## 7. Author Approval

All authors critically revised the manuscript and approved the final submitted manuscript.

## 8. Data Availability

For data privacy reasons, the raw data used for this study cannot be published. The figures within the article and supplementary material show detailed analyses for all patients used. Requests to access these datasets should be directed to: Deborah Nairn.

## Acknowledgements

We gratefully acknowledge financial support by Deutsche Forschungsgemeinschaft (DFG) through DO637/22-3, by the Ministerium für Wissenschaft, Forschung und Kunst Baden-Württemberg through the Research Seed Capital (RiSC) program and by Medtronic.

## Notes

**Conflict of Interest Statement:** RF, BF and AC are employees of Adas 3D Medical. This investigator-initiated study was financially supported by Medtronic. Medtronic had no influence on collection, analysis and interpretation of data, in the writing of the report, and in the decision to submit the article for publication.

### Competing Interest Statement

RF, BF and AC are employees of Adas 3D Medical. This investigator-initiated study was financially supported by Medtronic. Medtronic had no influence on collection, analysis and interpretation of data, in the writing of the report, and in the decision to submit the article for publication.

### Author Declarations

From February to July 2019, 41 consecutive patients with symptomatic persistent AF (lasting &gt;7 days and &lt;12 months) scheduled for their first PVI were prospectively included in this clinical trial (German WHO primary registry DRKS, unique identifier: DRKS00014687). The study was approved by the institutional ethics committee of the University of Freiburg (Germany) and all patients provided written informed consent prior to enrolment.

