## Supplementary Material for "LGE-MRI for diagnosis of left atrial cardiomyopathy as identified in high-definition endocardial voltage and conduction velocity mapping"

### 1. Supplementary Methods

#### *1.1. Electro-anatomical mapping*

Bandpass filtering at 16-500 Hz was applied to all bipolar electrograms using the CARTO-3 system. Any electrograms obtained when the electrodes were >7 mm from the atrial surface were removed from the analysis. Manual quality assessment was conducted to discard electrograms containing only noise or major artefacts. The system calculated the bipolar voltage values as the peak-to-peak amplitude of a single atrial beat in SR (the window of interest was restricted to the PR-interval with exclusion of the QRS complex). In this window, the local activation times (LAT) were also determined automatically by locating the maximum amplitude of the bipolar signal. The LAT was adjusted manually if it was incorrectly detected. Areas demonstrating LVS were confirmed using a separate contact force-sensing mapping catheter with a contact threshold of >5 g.

The CV was calculated from the LAT maps using the algorithms based on the polynomial fit technique described by Nagel et al. [1]. A feature to detect areas of potential wavefront collisions was additionally used. In these areas, the CV was recalculated in a second iteration in regions where only homogeneous excitation propagation occurred. The range of CV values can vary depending on the density of the fibrosis, which is unknown in this study [2].

#### *1.2. Late gadolinium enhancement magnetic resonance imaging (LGE-MRI)*

##### *1.2.1. Image intensity ratio method*

For the IIR method, a mid-myocardial layer was defined to create a shell in 3-dimensional space [3]. Left atrial mean blood pool intensity was automatically identified and IIR was calculated as the ratio between signal intensity of each voxel of the mid-myocardial LA layer normalized by the mean blood pool intensity. As described by Benito et al., voxels with an IIR of >1.20 were considered as pathological substrate and an IIR of >1.32 as dense scar [3].

##### *1.2.2. Utah method*

For the Utah LGE segmentation method, endocardial and epicardial borders of the LA were manually traced. Subsequently, the threshold was calculated as 2 to 4 standard deviations above the mean of the “normal tissue” on a slice-by-slice basis based on the operator’s opinion [4, 5].

#### *1.3. LA mean geometry*

A mean LA geometry built for a previous study was used [6]. The geometries obtained from each modality for all 36 patients were then aligned to the mean LA shape automatically using the iterative closest point algorithm with Scalismo, a statistical shape modelling software [7, 8]. After, Gaussian process morphable models were used to obtain a dense correspondence between each geometry and the mean LA. This is the same approach as used for building statistical shape models [9]. This approach yields the same number of surface points representing the same anatomical landmarks for each geometry. In this way, the patient's voltage, CV, LGE-MRI intensity values and Utah classification could be obtained for every point of the mean geometry.

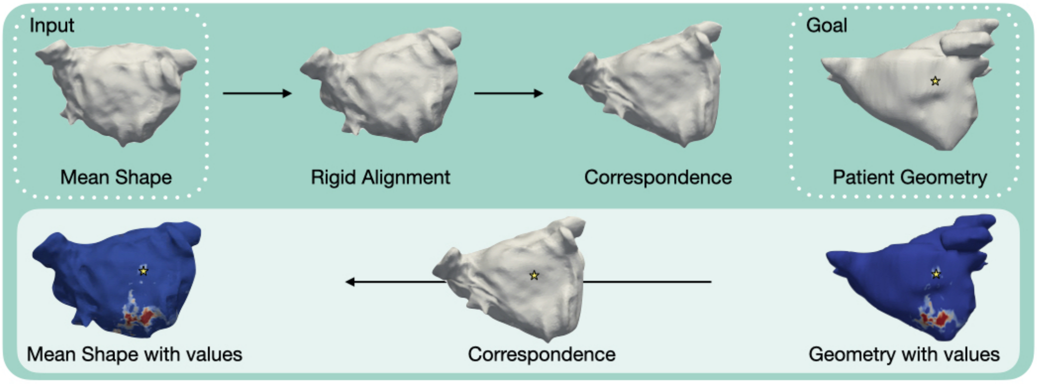


Figure 1: **Schematic diagram of the patient's information being projected onto the mean shape for an example patient.** On the top row, the steps applied to the mean shape to find correspondence with the patient-specific geometry are shown. The bottom row illustrates that a point is chosen (the star) and identified on the correspondence map, which has the same point ID's as the mean shape so that the information can be transferred back.

#### *1.4. Region-specific analysis*

The regions were obtained semi-automatically: first, rings around the PVs, MV and the left atrial appendage (LAA) were drawn using Blender, a 3D computer graphics software [10]. Following, the Eikonal equation was solved to obtain the shortest path between all rings. Paths were then chosen, which split the atria into the aforementioned regions.

### 2. Supplementary Results

#### *2.1. Patient characteristics*

| Patient Characteristics | Total = 36 |
|---|---|
| Age (years) | 66 ± 9 |
| Male (%) | 31 (84) |
| BMI ($kg/m^2$) | 27.4 ± 3.5 |
| LVEF (%) | 57 ± 8 |
| LA diameter (AP, mm) | 47 ± 6 |
| $CHA_2DS_2$-VASc score | 2 (1-3) |
| Hypertension (%) | 27 (73) |
| Diabetes mellitus (%) | 3 (8) |
| Prior stroke or TIA (%) | 1 (3) |
| Coronary artery disease (%) | 5 (14) |
| Procedure duration (min) | 146 ± 19 |
| Successful circumferential PVI (%) | 36 (100) |
| CTI ablations (%) | 8 (22) |
| Prior antiarrhythmic therapy (%) | 29 (78) |

Table 1: **Patient clinical demographics.** Abbreviations: BMI = body mass index, LVEF = left ventricular ejection fraction, LA = left atrial, AP = anterior-posterior, TIA = transient ischemic attack, PVI = pulmonary vein isolation, CTI = cavotricuspid isthmus .

| Electro-anatomical Mapping | SR |
|---|---|
| Map points (pts) | 2098 ± 515 |
| Bipolar voltage (mV) | 1.84 ± 1.59 |
| CV (m/s) | 0.48 ± 0.49 |
| LVS surface area, SR < 0.5 mV ($cm^2$(%)) | 14 ± 19 (7 ± 10) |
| Slow CV surface area, CV < 0.5 mV ($cm^2$(%)) | 22 ± 24 (12 ± 13) |

| LGE-MRI | |
|---|---|
| IIR Intensity values | 1.01 ± 0.23 |
| High intensity surface area, IIR > 1.2 (cm2 (%)) | 37 ± 27 (20 ± 15) |
| UTAH fibrotic surface area ($cm^2$(%)) | 22 ± 10 (12 ± 5) |

Table 2: **Mapping information of patients included in the study.**

### 2.2. Spatial distribution of electro-anatomical and LGE-MRI maps

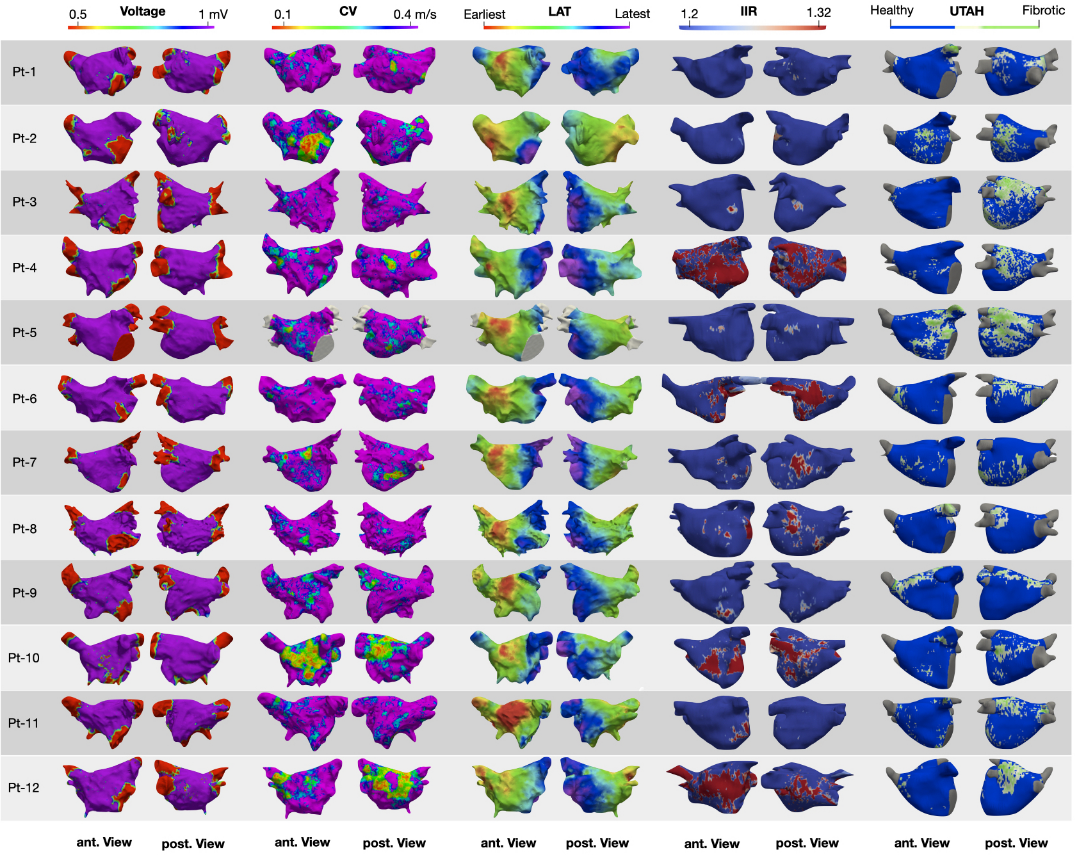


(a) Patient 1-12

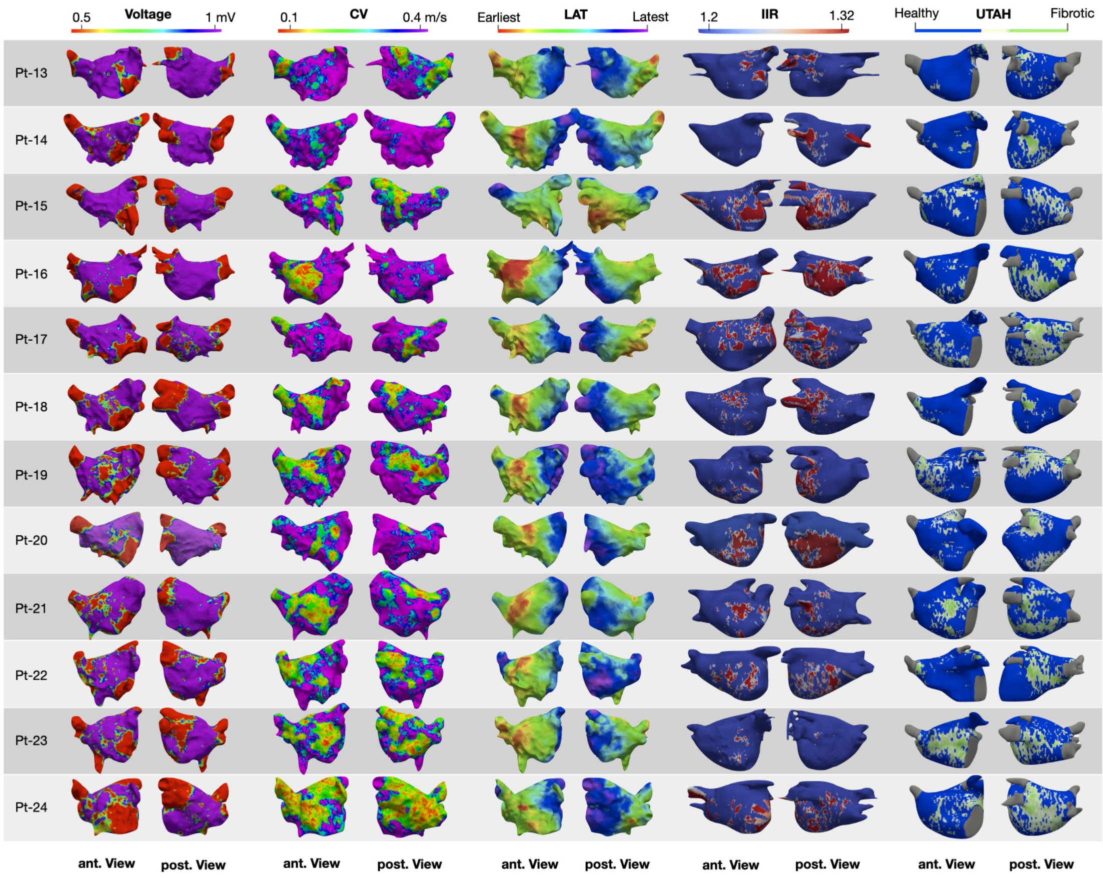


(b) Patient 13-24

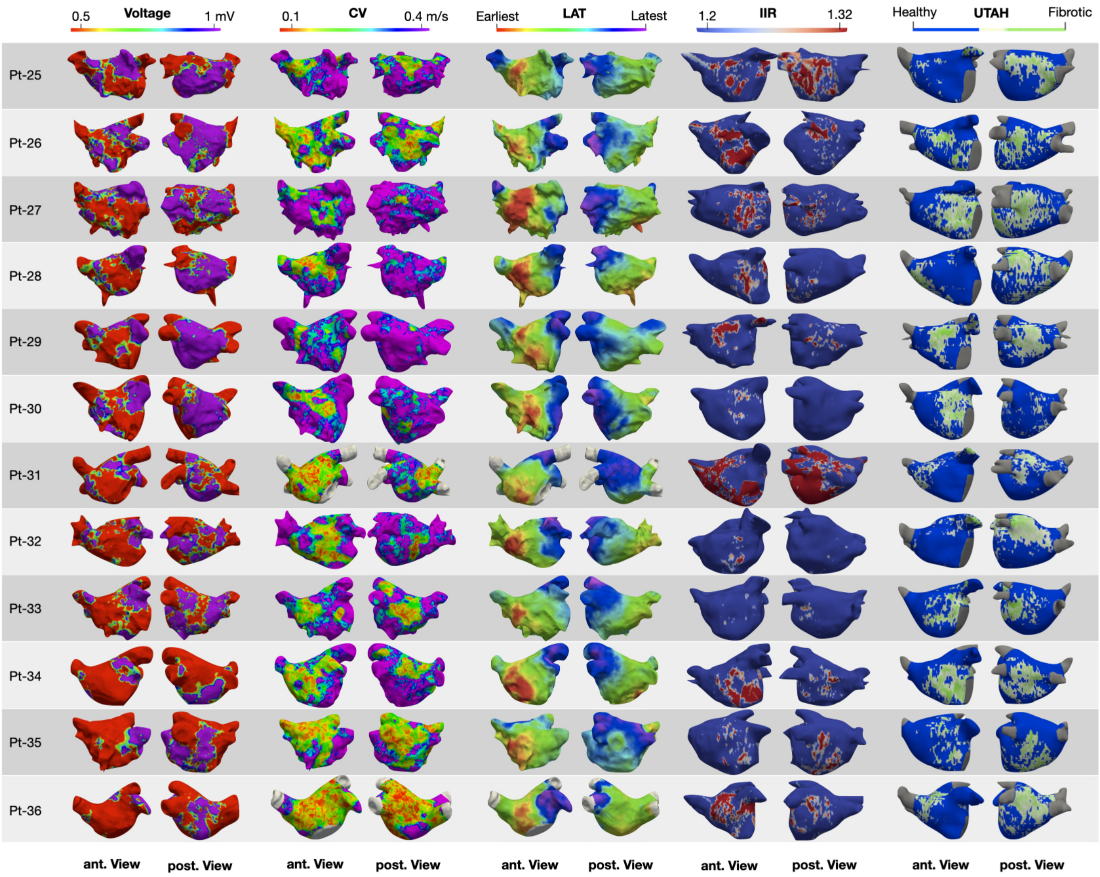


(c) Patient 25-36

Figure 2: **Three-dimensional distribution patterns of the pathological substrate in different mapping modalities for all patients.** Each row shows one patient and each column pair shows the anterior and posterior view for that patient. On the left the voltage map can be seen, with voltages below 0.5 mV in red and above 1 mV in purple. Next the conduction velocity between 0.2 and 0.4 m/s. The LAT map is then shown from earliest to latest activation for each patient. Following is the LGE-MRI using the IIR method shown with cut off values between 1.2 and 1.32. Finally, the LGE-MRI using the Utah method is shown, with fibrotic areas in green and healthy areas in blue.LVS and slow CV was found to extensively cover the anteroseptal wall in all patients presenting LVS ≥5% of total LA at <0.5 mV.

#### 2.3. Optimising the LGE-MRI threshold to identify LVS

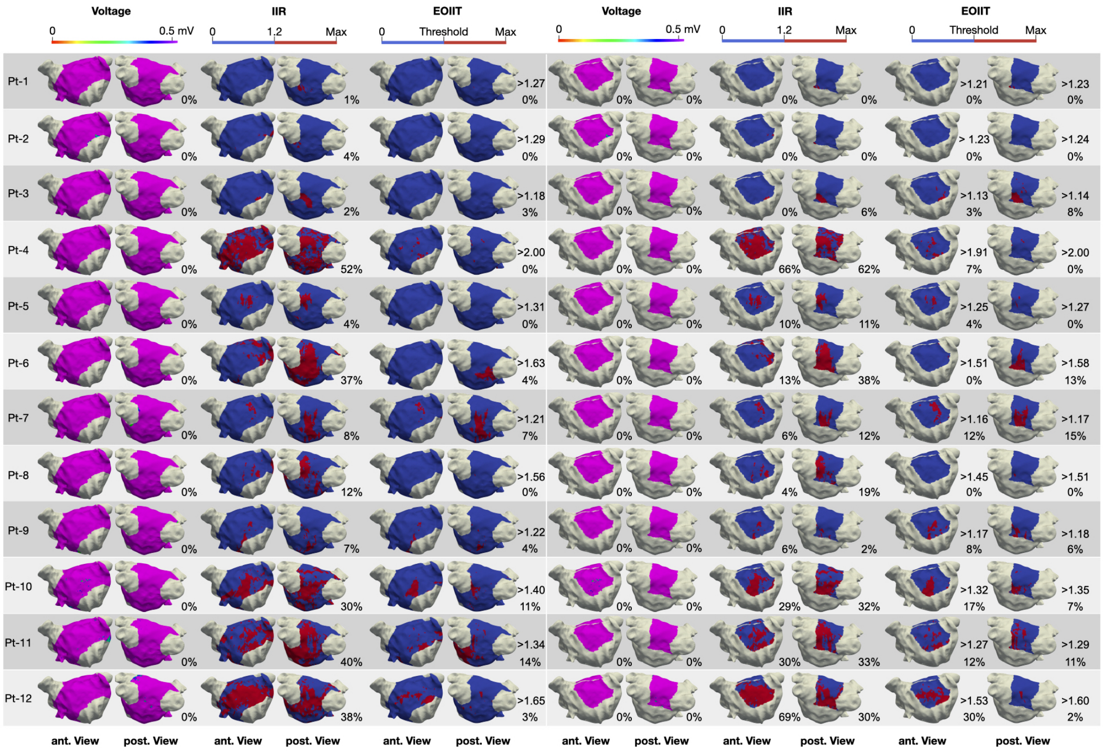

(a) Patient 1-12

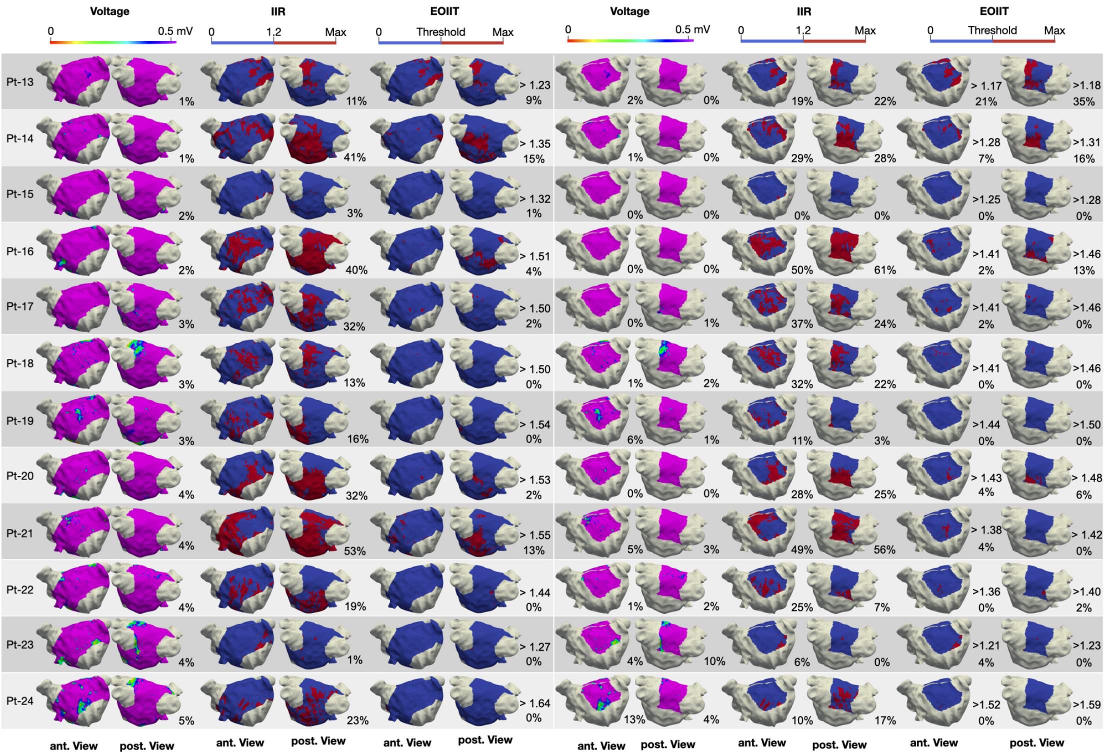


(b) Patient 13-24

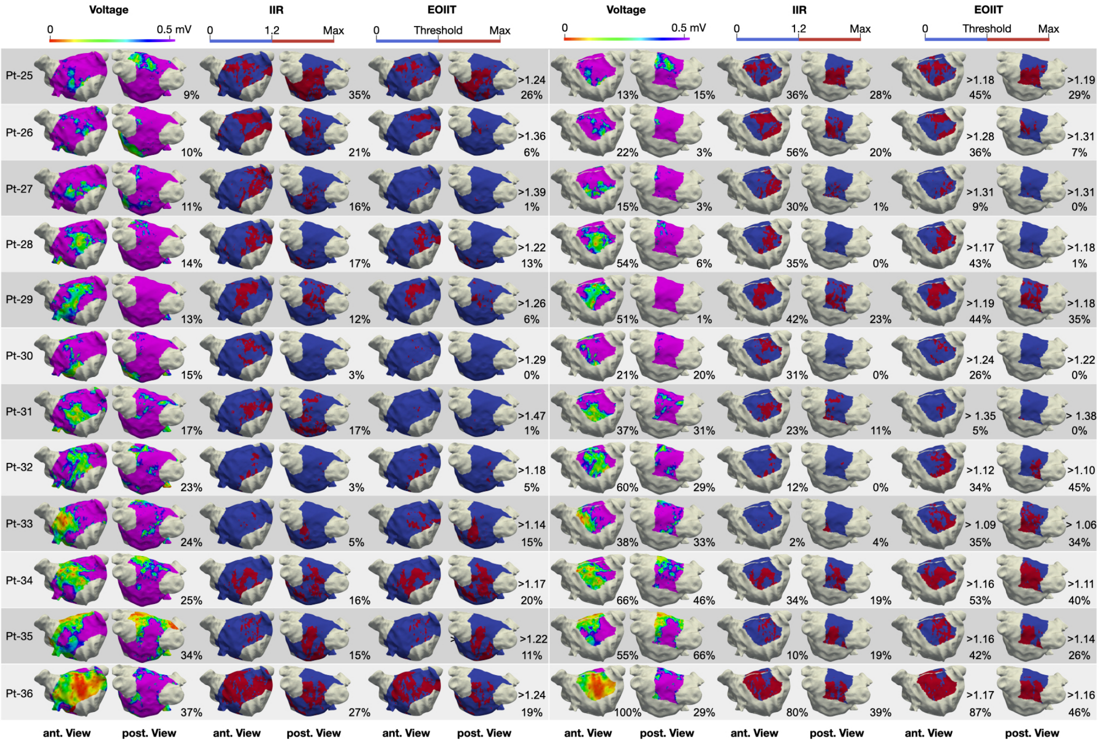


(c) Patient 25-36

Figure 3: **Three-dimensional voltage and LGE-MRI maps showing the high intensity areas defined by the IIR ¿ 1.2 threshold and using the new EOIIT thresholds.** Each row shows one patient and each column pair shows the anterior and posterior view for that patient. On the left hand side the percentage of low voltage and high intensity can be seen for the entire atria excluding the PV's and MV. On the right, the percentage can be seen for the the anterior and posterior wall seperately. The new EOIIT threshold for each patient is given divided by that patients mean bloodpool value to allow for quick comparability to the IIR method.

#### *2.4. Optimising the LGE-MRI threshold to identify low-voltage substrate*

To quantify the performance of the new EOIIT, the percentage of LGE-MRI extent defined by the new thresholds was plotted against the percentage of LVS extent (<0.5 mV) as shown in figure 4. Figure 4 shows for the anterior wall, a substantial improvement (y = 0.63x + 5.6, $R^2$ = 0.68, p <0.001) occurred when using the new EOIIT compared to the IIR >1.2 threshold (y = 0.28x + 21.3, $R^2$ = 0.11) with the optimal relationship being a diagonal line. From figure 4 the Bland-Altman plot shows that previously the LGE-MRI was measuring on average 10% more pathological substrate than the voltage map. Using the new thresholds, this value is reduced to 0%. Figure 5 shows the plots for the entire atrium and the posterior wall. Only a slight improvement was seen in estimating the percentage of LV extent from the MRI compared to using the IIR >1.2 threshold. Additionally, the new EOIIT thresholds show an improvement (y = 0.56x + 13.9, $R^2$ = 0.22) in identifying the same extent of substrate as when assessing slow conduction areas compared to using the IIR 1.2 threshold 6. However, the best relationship is seen with CV and LVS (y = 1.02x+9.7, $R^2$ = 0.57).

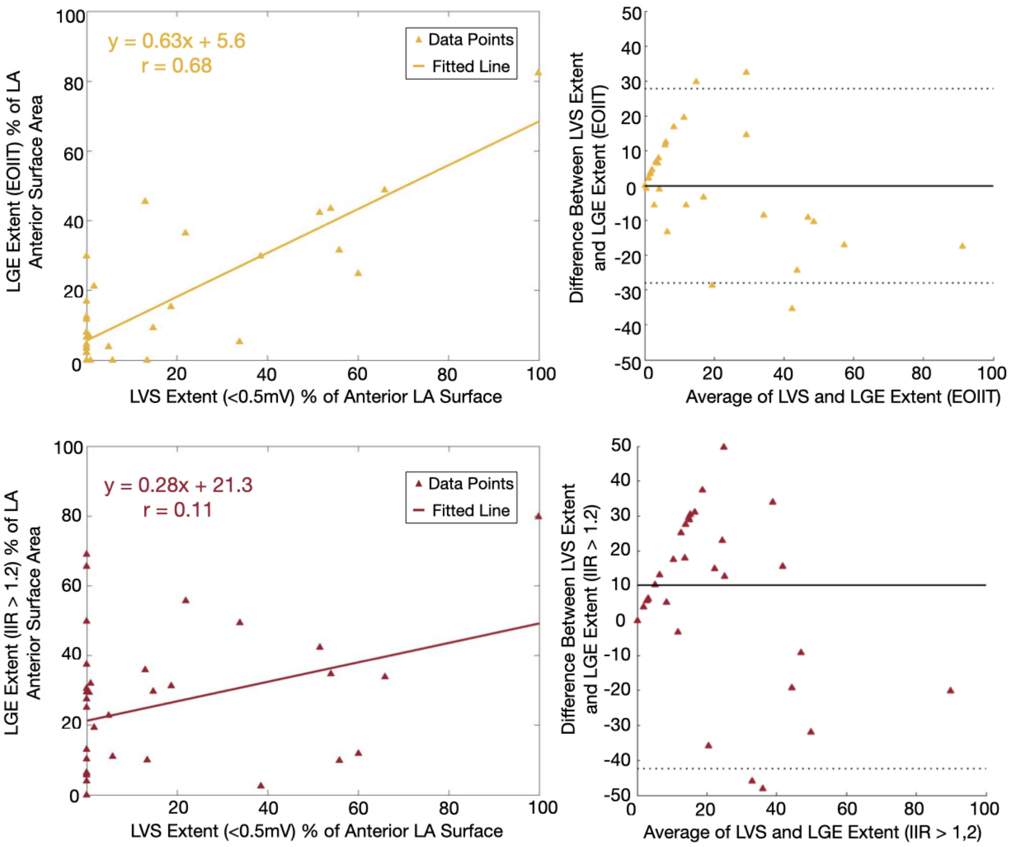


Figure 4: **Percentage of LGE-MRI extent against the percentage of LVS extent and Bland-Altman plot for the anterior wall.** Each triangle represents one patient. On the top row, the LGE-MRI extent obtained using the new thresholds is shown. On the bottom row, the extent is based on IIR >1.2.

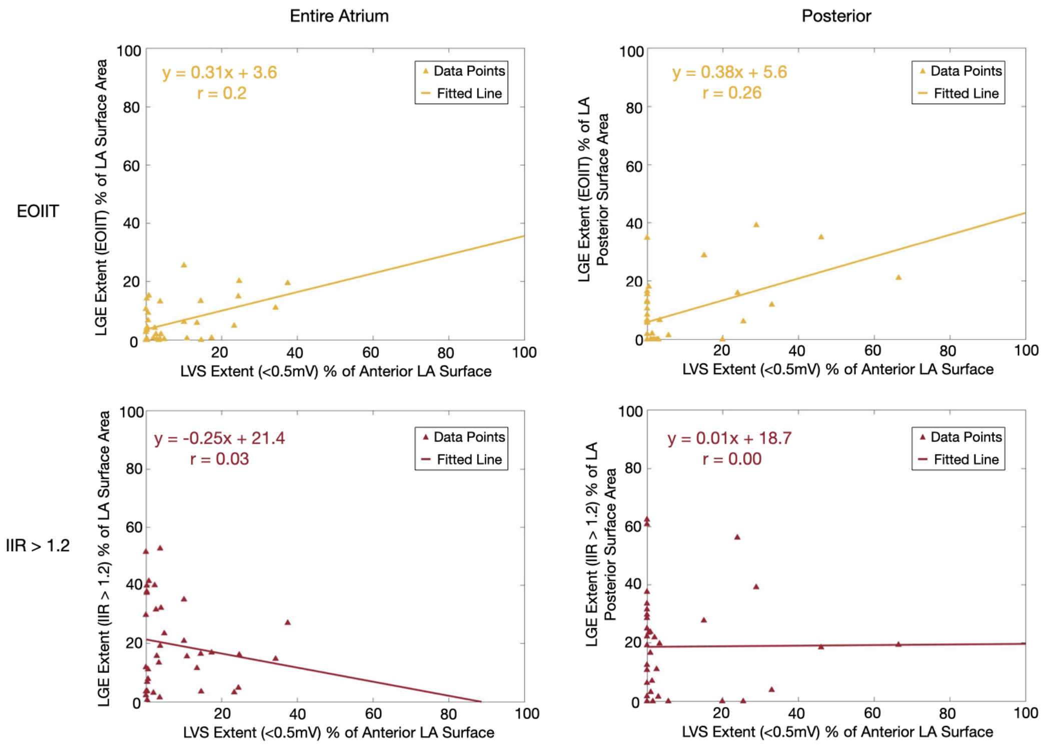


Figure 5: **Percentage of LGE-MRI extent against the percentage of LVS extent for the entire atrium and posterior wall.** Each triangle represents one patient. On the top row, the LGE-MRI extent obtained using the new thresholds is shown. On the bottom row, the extent is based on IIR >1.2.

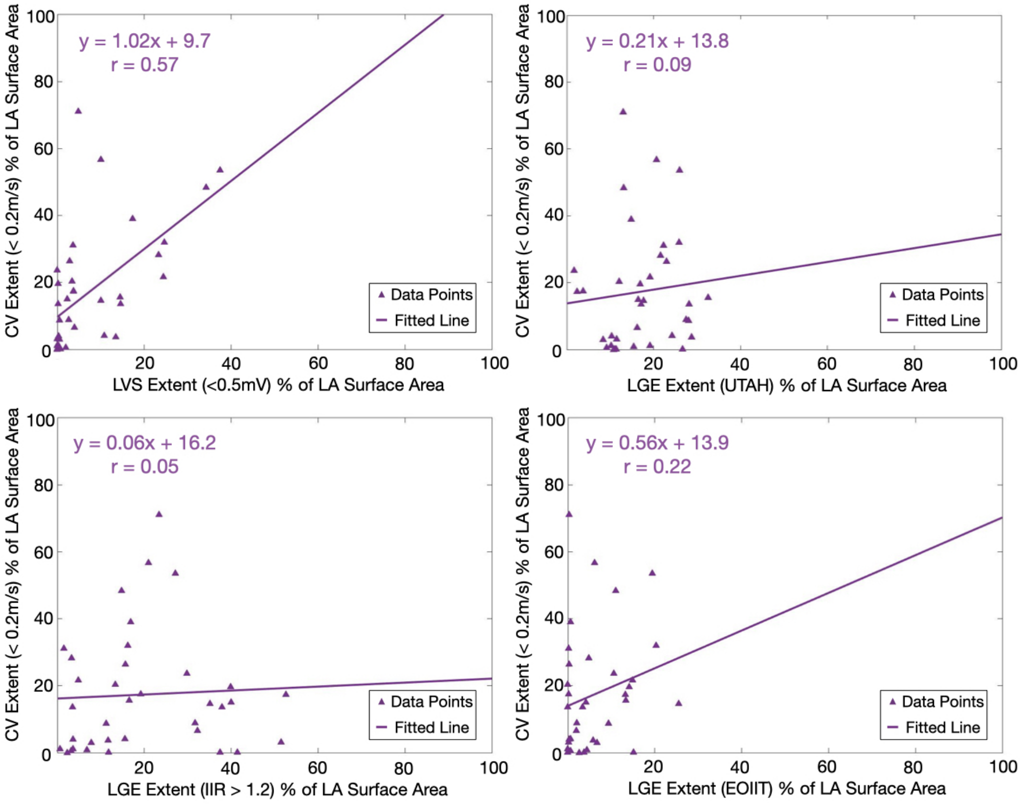


Figure 6: **Percentage of CV <0.2 m/s extent against the percentage of LVS <0.5 mV (top left), LGE-MRI UTAH (top right), LGE-MRI IIR >1.2 (bottom left) and EOIIT (bottom right) extent for the entire atrium.** Each triangle represents one patient.
